# Exploring the diagnostic and prognostic utility of resting state functional MRI connectivity in acute/sub-acute TBI

**DOI:** 10.1101/2025.05.20.25327881

**Authors:** M. K. Chepisheva, X. Shen, C. Lacadie, W. Luo, J. Appleton, J. Arora, J. Bhawnani, A. Mahajan, B. Omay, E. Gilmore, B. Edlow, R. T. Constable, J. A. Kim

## Abstract

Traumatic brain injury (TBI) is a leading cause of disability worldwide. Yet, our understanding of the mechanisms of this condition is limited, especially in the acute setting. Clinical assessments such as the Glasgow Coma Scale (GCS) and modified Rankin Scale (mRS) are widely adopted due to their practicality, despite limited specificity for underlying network disruption. Here, we examined the relationship between resting-state functional connectivity (FC) and admission GCS and 3-month mRS to assess whether FC provides network-level representation of brain dysfunction captured within the limits of these standard clinical measures.

We performed a retrospective analysis of resting-state functional MRI and clinical data in 58 patients (41.28 ± 18.63) scanned acutely/subacutely (≤ 31 days). Then, for a secondary analysis, we included 50 more patients who presented after either a first or a repeat incident and were scanned either acutely/subacutely or chronically (<2 yrs) (altogether 108 patients, 46.4 ± 20.1yrs). Using a 268-node functional atlas, we derived 35,778 unique edges, based on which we calculated the mean FC of 10 resting-state networks and used those to establish a link to TBI severity and functional outcome.

Our analysis showed that when dividing sub/acute patients (n=58) based on GCS severity, only the Subcortical network showed a significant discrimination between mild and moderate-severe GCS at admission (P<0.001), with hyperconnectivity noted in mild patients, and hypoconnectivity in moderate-severe patients. This difference appeared to be driven by the thalami (Right, P=0.002; Left, P<0.001). Similar results were observed when investigating GCS subscores at admission (Eyes, Motor, Verbal, all P<0.001). Further, when evaluating mRS outcomes at 3-months against FC, differences were noted within the Motor, Cerebellum and Medial-Frontal networks, though none survived multiple comparisons. Importantly, we found the DMN and mRS to be correlated but with a limited relationship (r^2^= 0.18). Lastly, we performed a post-hoc analysis (n=108) to investigate if the hyperconnectivity in the Subcortical network of sub/acute mild GCS patients remained irrespective of acuity of scanning or frequency of TBI. Our analysis showed that GCS severity appeared to be the main driver of FC within the Subcortical network, whereas acuity of scanning, alongside GCS severity, contributed to the results of chronically scanned patients.

While GCS and 3-month mRS scores offer some meaningful insights, their limited capture of the neural representation underscores the need to investigate whether other early clinical assessments correlate more robustly with early resting-state networks or if such networks themselves could predict future outcomes.

## Introduction

Traumatic brain injury (TBI) is a leading cause of disability worldwide. Yet, our understanding of the pathophysiology and mechanisms of this condition across the full spectrum of injury remains incomplete. TBI severity has long been defined by clinical rating scores, of which the Glasgow Coma Scale (GCS) ^1^ has been the most commonly used. This is due to its widespread acceptance, ease of application and historical precedence. However, the reliability of GCS as a defining feature of TBI acuity has been increasingly debated ^2, 3, 4, 5^. Cited reasons include observer experience, subjectivity in assessing components of the GCS, demographic differences (e.g., language barriers), toxin or sedation effects, different evaluation settings, treatment factors (e.g., intubation), injury factors (e.g., spinal cord or orthopedic injury limitations) ^5^. Thus, groups have begun to propose new frameworks to more wholistically characterize acute TBI^6^. Similarly, functional outcomes in TBI have been limited in the clinical setting and their reliability needs to be better defined ^7^, as potential interobserver variability might be of concern ^7^.

Clinicians and researchers alike have sought to refine the various characteristics which encompass the heterogeneous nature of TBI as improved knowledge about its mechanisms would lead to an improved diagnosis, quality of life and outcome for the patients ^8, 9, 10^. Advanced neuroimaging, such as functional MRI, has become increasingly utilized to study TBI patients ^11, 12, 13^. However, its application remains primarily limited to the research setting or small-scale clinical pilot studies. In the clinical setting, structural MRI has been used to identify lesion burden and predict recovery after TBI, but the additional diagnostic and prognostic utility of more advanced MRI sequences is less well-defined. More recently, resting-state functional MRI (rsfMRI) has been employed to investigate whether alterations in the brain’s intrinsic connectivity patterns reflect TBI pathophysiology, with some studies reporting impaired small world topology and reduced network efficiency post TBI ^14, 15, 16^.

Despite these advancements, establishing a functional neuroimaging-based biomarker of TBI severity, especially in the early stage of TBI, remains a challenge. Many rsfMRI studies are focused on chronic TBI patients and reported incomplete recovery and persistent symptoms, such as cognitive impairment, apathy or fatigue, all of which stem from persisting regional or global changes in the brain’s architecture ^17, 18, 19^. Even patients with mild TBI can report symptoms up to 12 months post injury, and those with moderate-severe TBI can experience symptoms for an even longer duration ^20, 21^. Prior studies report multiple regions with both increased and decreased FC (FC), likely due to: (1) heterogeneous TBI etiologies, (2) varied brain regions under investigation, (3) studies spanning the full spectrum of TBI severity (mild, moderate, severe) and (4) differences in stage (acute, chronic) at the time of acquisition. The high variability that has been generally observed in the field ^22, 23^ and the large number of prognostic models ^11, 24, 25, 26, 27^ impede a consensus regarding possible outcome biomarkers of TBI.

To elucidate this knowledge, more real-world studies are needed. Despite their potential benefits ^28^, rsfMRI studies in the clinical setting are rare due to patient acuity of illness and limited ability to tolerate scans during the acute/sub-acute period post TBI. In the few studies which successfully acquired data in such a setting, research suggests that intact DMN connectivity is correlated with recovery of consciousness after TBI ^29^ and other severe illnesses with prolonged coma states like COVID-19 ^12^. In another intensive care unit study ^30^ examining 25 acutely unresponsive patients, researchers showed that the visual and left frontoparietal networks play a critical role in the successful prediction of subsequent recovery. Consequently, a broader evaluation of the canonical networks was proposed as more insightful compared to focusing solely on the importance of the DMN ^31, 30^. However, whether resting-state networks can be used to predict long-term functional outcomes beyond recovery of consciousness after TBI remains to be determined.

In this study, we investigated whether early resting-state FC in acute/subacute TBI is associated with commonly used clinical measures of TBI severity (GCS at admission) and 3-month functional outcome(Modified Rankin Scale). While limited in their ability to capture specific cognitive or network-level dysfunction, these scales are widely used. Our aim was therefore to identify their underlying network-level brain correlates, rather than to endorse them as optimal endpoints. By doing so, we seek to enhance the biological interpretability of rsfMRI findings in TBI and to inform future studies that integrate more specific, domain-relevant outcome measures.

## Material and Methods

### Participants/recruitment/selection criteria

We retrospectively screened 1,053 neurotrauma patients (05/2018 – 09/2022) who were seen in the Emergency Room (ER) of Yale New Haven Hospital (New Haven, CT, USA). Our main inclusion criteria were: (1) patients ≥18 years old, with a first TBI, (2) had no neurological condition at baseline, (3) did not sustain another TBI within 3-months of their initial TBI encounter, (4) had rsfMRI ≤31 days post-TBI. (Fig.1). Patients meeting these criteria were used for our main analysis (n = 58, mean ± SD, 41.28 ± 18.63 years, 33% F), (Table 1). A post-hoc analysis was also conducted for patients with either a first or a repeated TBI and an initial scan ≤31 or >31 days after their accident (altogether 108 patients, (mean ± SD, 46.4 ± 20.1 years)). Lastly, we selected 113 healthy controls (mean ± SD, 32.5 ± 12.6 years, 52% F) from a previously established cohort, described elsewhere ^32^, to establish a non-statistical comparison group for all figures. This study was conducted in accordance with the principles of the Declaration of Helsinki and approved by the IRB of Yale University School of Medicine.

**Figure 1.**
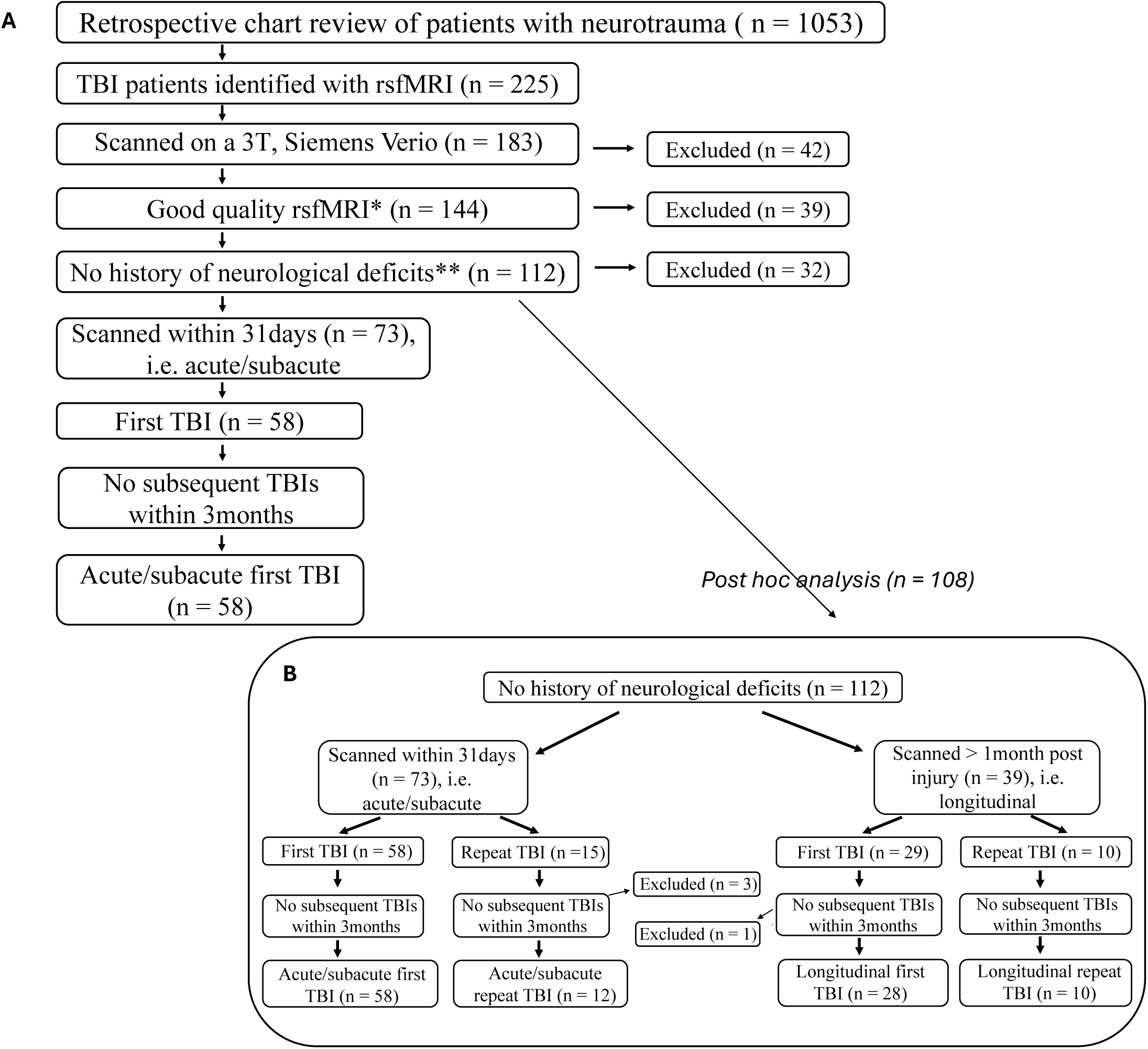
Study Flowchart of Participant Inclusion and Exclusion criteria: **(A).** Cohort of acute/subacute first TBI patients for primary analyses, based on the following inclusion criteria: (1) TBI patients ≥18 years old, who experienced their first TBI, (2) scanned within 31 days of their TBI event, (3) Good quality rsfMRI scans*, (4) no subsequent TBIs within 3months and (5) with no history of neurological deficit at baseline**. These selection criteria resulted in a final cohort of 58 TBI patients. **<u>(B)</u>.** Cohort for post-hoc investigation of the Subcortical network. The selection criteria included patients scanned acutely/subacutely and those with a first/(non-first = repeat) TBI. We identified 108 TBI patients and divided them into four groups: **(1)** sub-acute/acute first TBI (n = 58) - main cohort presented in Fig.1. A – patients who were scanned acutely/sub-acutely (i.e. <= 31 days) and had their first TBI, **(2)** sub-acute/acute repeat TBI (n = 12) – patients who were scanned acutely/sub-acutely (i.e. <= 31 days) but had a prior history of a TBI, **(3)** longitudinal first TBI (n = 28) – patients scanned > 31 days up to 2 years after their initial TBI encounter, with no prior history of TBI and **(4)** longitudinal repeat TBI (n = 10) – patients scanned > 31 days up to 2 years after the TBI event of interest with a history of a prior TBI. **<u>Key</u>:** rsfMRI = resting-state fMRI; **\***Good quality rsfMRI = was considered acceptable quality for resting-state analysis, i.e. frame-to-frame displacement of the functional image < 1.2mm, alongside no visual disturbances (e.g. lines) or other artifacts masking the brain image; **\*\***No history of neurological deficit at baseline = patients’ electronic medical records were examined to ensure no patients with previous history of a general neurological disorder/brain condition (e.g., brain cancer, stroke, epilepsy, bipolar disorder), neurodegenerative disorder (e.g., Huntington’s disease, mild cognitive impairment (MCI) or dementia (e.g., Alzheimer’s disease (AD), fronto-temporal dementia (FTD) or vascular dementia (VaD))) or neurodevelopmental disorder (e.g., Down syndrome, Social Anxiety Disorder (SAD)) were included in the study.; Excluded patients were those whose conditions did not comply with the general requirements.

**Table 1.** Demographics of Acute/Subacute First TBI patients (Main Cohort, n = 58). Columns present information about the total cohort (n = 58), the mild GCS patients (n = 28) and the moderate-severe (n = 30). **<u>Key</u>:** GCS=Glasgow Coma Scale; GCS subscores = GCS Eyes + GCS Motor + GCS Verbal; mRS = Modified Ranking Scale; Mechanism of Injury (MOI) - accel/decel = acceleration/deceleration, fall > 3f = fall from a higher elevation, fall from standing = fall from a standing position or a normal walking position, direct impact = indicates a hit to the head by an object, not clear – no clear etiology of TBI.

|  | **Total Cohort** | **Mild GCS Subgroup** | **Moderate-severe GCS Subgroup** |
| --- | --- | --- | --- |
| *Sample size (n)* | 58/58 | 28/58 | 30/58 |
| *GCS at admission (Mdn [IQR])* | 10.5 [7,15] | 15 [14.75, 15] | 7 [6,8.75] |
| *Eyes* | 3 [1,4] | 4 [4,4] | 1 [1,2] |
| *Motor* | 6 [4,6] | 6 [6,6] | 4 [4,5] |
| *Verbal* | 3.5 [1,5] | 5 [5,5] | 1 [1,1.75] |
| *mRS at 3months (Mdn [IQR])* | 2 [1,3] | 1 [1,2] | 3 [2,4] |
| *Age (Mean ±SD)* | 41.28 ± 18.63 | 48.29 ± 19.85 | 34.73 ± 14.97 |
| *Gender, % F* | 33% | 43% | 23% |
| *Mechanism of injury, n(%/within group)* |  |  |  |
| *accel/decel* | 39 (67%) | 12(43%) | 27(90%) |
| *fall > 3f* | 5 (9%) | 5(18%) | - |
| *fall from standing* | 11 (19%) | 10 (36%) | 1(3%) |
| *direct impact* | 2(3%) | - | 2(7%) |
| *not clear* | 1 (2%) | 1(3%) | - |

### Clinical assessment

#### Glasgow Coma Score (GCS)

GCS is a standard clinical assessment ^1^ comprised of three separate subscores making up a total score. Clinical staff assesses patients’ Eyes opening (scored from 1 - 4), Motor (1 - 6) and Verbal (1 - 5) responses. These subscores were calculated based on clinical notes at the time of admission and summed to produce a total GCS score. The latter ranges from three (no eye opening/ motor/verbal response) to 15 (fully alert/oriented/following commands). In this study, GCS values at admission were extracted from electronic health records and adjudicated by a neurointensivist (JAK).

#### Modified Rankin Scale (mRS)

The mRS ^33, 34^ measures the degree of disability in patients following a stroke. It uses a 7-degree scale (0-6), where 0 indicates “no symptoms” and 6 – death. In a TBI cohort, GOS-E (Glasgow Outcome Scale – Extended) or GOS is more typically used. However, many functional outcomes measures used in TBI, including those listed, have similar validity concerns ^35, 36, 37^. Additionally, this was a retrospective study limited by the availability of information from clinical health records. This precluded accurate GOS-E calculation. Given GOS is a coarser scaling than mRS with an otherwise strong agreement,^38, 39,40^ (Supplementary Fig.2), we chose mRS as our outcome evaluation. All mRS values were extracted from the latest patient note and adjudicated by a neurointensivist (JAK).

**Figure 2.**
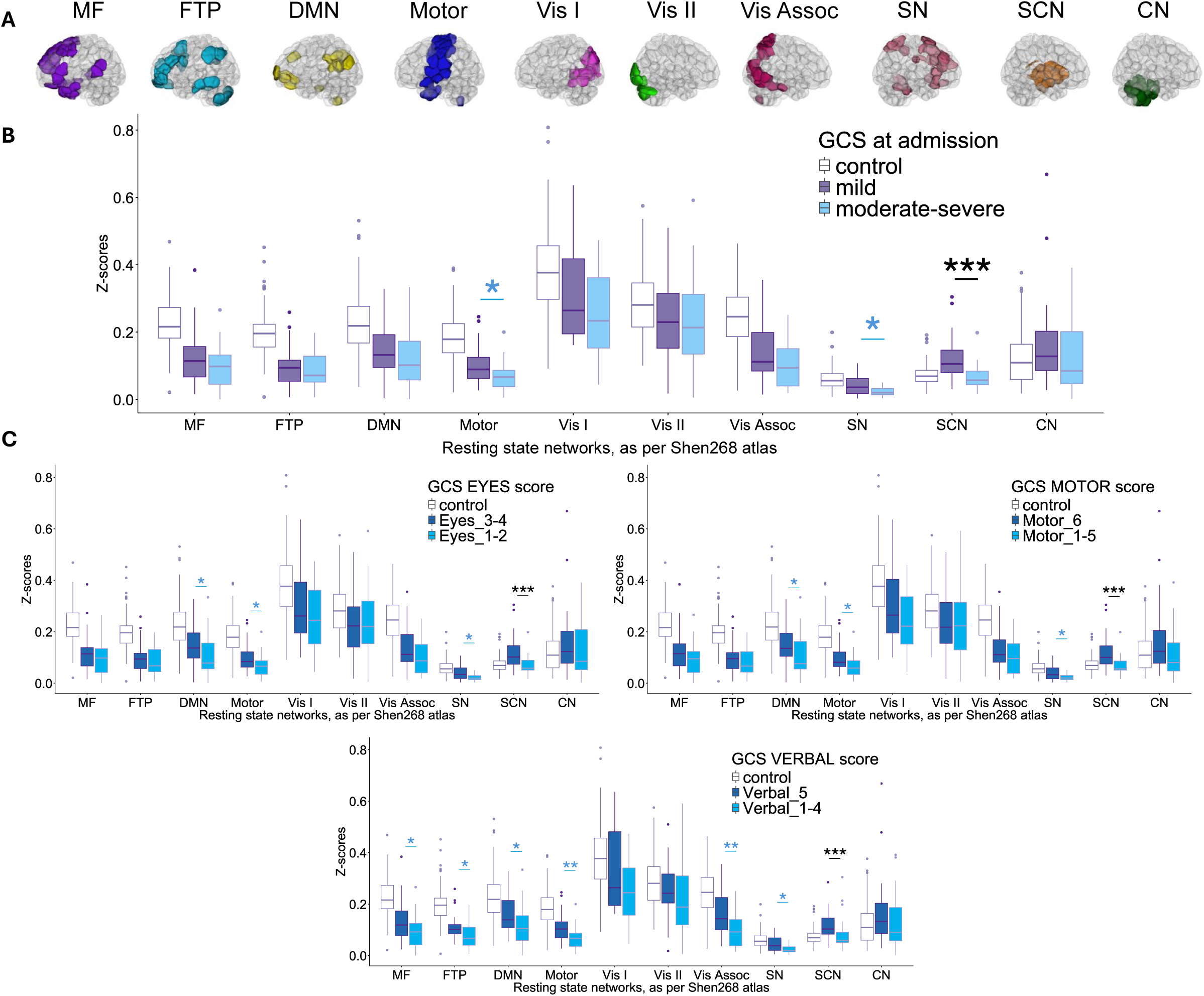
Association between Glasgow Coma Scale (GCS) and functional connectivity across 10 resting-state networks. **(A).** Ten resting-state networks derived from the Shen*268* functional atlas ^42^. These are Medial-Frontal (MF), Fronto-Parietal (FTP), DMN (Default Mode Network), Motor, Visual I (Vis I), Visual II (Vis II), Visual Association (Vis Assoc), Salience (SN), Subcortical (SCN) and Cerebellum (CN) networks, respectively ^42,43,44,40^.This image was prepared using an In-house Software - Yale BioImage Suite (https://bioimagesuiteweb.github.io/webapp/connviewer.html?species=human). **(<u>B)</u>.** Network analysis based on TBI severity classified by Total GCS on admission score (moderate-severe patients (GCS 3-12, n = 30) vs mild (GCS 13-15, n = 28)). The Mann-Whitney U test (total sample size n = 58, divided into n(mild) = 28 and n(moderate-severe) = 30) was used to establish any association between the Z-score connectivity maps of the resting-state networks and patients’ GCS severity (i.e. moderate-severe vs mild). The x-axis displays the 10 resting-state networks based on the Shen*268* functional atlas, and the y-axis shows the functional connectivity Z-scores. Healthy controls (n = 113) are displayed for visual reference only. Each single point on the figure represents the Z-score of an individual participant’s functional connectivity map (i.e. patient or control) for the corresponding resting state network. **(<u>C)</u>.** Network analysis for each GCS subscore on admission. Each GCS subscore was dichotomized: <u>GCS Eyes Subscore</u>: 1-2 vs 3-4 (1 = no response, 4 =spontaneous eye opening); <u>GCS Motor Subscore</u>: 1-5 vs 6 (1 = no response, 6 = obeys command); <u>GCS Verbal Subscore</u>: 1-4 vs 5 (1 = no response, 5 =fully oriented). The Mann-Whitney U test (total sample size n = 58, divided into EYES [n(scores_3-4) = 32, n(scores_1-2) = 26], MOTOR [n(score_6) = 33, n(scores_1-5) = 25] and VERBAL[n(score_5) = 23, n(scores_1-4) = 35]) was used to establish any association between the Z-score connectivity of the resting-state networks and patients’ GCS subscores (i.e. Eyes, Motor and Verbal scores, respectively). The x-axis displays the 10 resting-state networks based on the Shen*268* functional atlas, and the y-axis shows the functional connectivity Z-scores. Healthy controls (n = 113) are displayed for visual reference only. Each single point on the figure represents the Z-score of an individual participant’s functional connectivity map (i.e. patient or control) for the corresponding resting state network. **<u>Key:</u>** blue * denotes significant results that did not survive multiple comparisons; black * are results that remain statistically significant after Bonferroni corrections, * *p* < 0.05, ** *p* < 0.01, *** *p* < 0.001; Medial-Frontal network = MF, Fronto-Parietal network = FTP; Default Mode Network = DMN, Visual I network = Vis I, Visual II network = Vis II, Visual Association = Vis Assoc, Salience = SN, Subcortical network = SCN, Cerebellar network = CN.

### Imaging data

#### TBI patients

Structural and functional (rsfMRI) imaging was performed on a 3T Siemens Magnetom Verio Syngo clinical scanner at Yale New Haven Hospital. The TBI protocol used for the resting-state acquisition was with the following parameters: number of slices = 34, FoV = 220mm, repetition time (TR) = 1760 ms, echo time (TE) = 25 ms, flip angle = 78°, matrix size = 64 × 64 (voxel size = 3.4 x 3.4 x 3.4mm), in-plane resolution = 3.4 mm^2^, slice thickness = 3.4 mm. Parameters of the anatomical magnetization were as follows: number of slices per slab = 176, FoV = 250mm, TR = 2300 ms, TE = 2.34 ms, flip angle = 8°, matrix size = 256 × 256 (voxel size = 1.0 x 1.0 x 1.0mm). The duration of the resting-state acquisition was 9 mins, with no specific instructions given to the patient.

#### Healthy Controls

Structural and resting-state FC data for the group of 113 healthy controls were used from a previously acquired dataset ^32^. This dataset was collected at the Yale Magnetic Resonance Research Center (MRRC, Yale University, New Haven, CT, USA) on a 3T Siemens Trio TIM scanner ^32^. The resting-state sequence was with the following parameters: repetition time (TR) = 1000 ms, echo time (TE) = 30 ms, flip angle = 62°, matrix size = 84 × 84, in-plane resolution = 2.5 mm^2^, slice thickness = 2.5 mm. The structural (MPRAGE) sequence had the following parameters: 176 sagittal slices, TR = 2530 ms, TE = 3.32 ms, flip angle = 7°, matrix size = 256 × 256, in-plane resolution = 1.0 mm^2^, slice thickness = 1.0 mm ^41^.

### Preprocessing

Anatomical and functional preprocessing was performed in Yale University’s BioImage Suite ^42^ using several custom scripts/functions in bash and Matlab (Mathworks). The standard procedure was described in detail elsewhere ^43, 44, 45^ and was applied to the TBI patients and healthy controls. In brief, it included skull-stripping, non-linear registration (for anatomical images) and slice-timing, motion correction (for functional images using SPM8 https://www.fil.ion.ucl.ac.uk/spm/software/spm8/), linear registration (anatomical and functional images), and calculation of the resting-state FC matrices using the Shen268-node brain atlas. Of note, there were additional procedures applied during the standard pre-processing, including regression of mean time-courses in white matter (WM), cerebrospinal fluid (CSF) and global signal regression (GSR) and low-pass Gaussian filtering (approximate cut-off frequency of 0.12 Hz) ^45^. Functional runs were discarded if the mean frame-to-frame displacement was greater than 1.2mm. The results after each step were individually inspected to ensure reliable pre-processing output.

Considering some brain regions are more susceptible to signal loss during functional MRI acquisition (e.g. air/tissue, bone/tissue), special care was taken to investigate the signal availability in all nodes of each subject. If any such “signal-loss” nodes were noted even in a single patient, we applied the conservative approach of excluding these nodes in all subjects ^44^. After the above pre-processing steps, only two such nodes were removed from all patients/controls individual connectivity matrices.

### Node and network definition

In this study, we used the functional Shen268-*node* atlas that was derived based on rsfMRI resting-state data from a cohort of healthy participants ^46, 47^. After the parcellation of the data into 268 nodes for each subject (i.e. patient/ control) ^44^, the mean time-course of each region of interest (i.e. a node) was computed. Then, the Pearson *r* correlation was calculated between each pair of nodes to yield a *268 × 268* matrix of correlations representing the function connections, or *edges* between nodes ^45, 41^. Importantly, the Pearson correlation coefficients were Fisher Z-transformed at the individual level. Considering only the upper triangle of the FC matrix, this calculation resulted in 35,778 unique edges for the whole brain ^45^.

Further, for the ROI analysis, the 268 nodes were assigned to 10 functionally derived resting-state networks ^46, 47, 48, 44^: (1) Cerebellum (CN), (2) DMN, (3) Fronto-parietal (FTP), (4) Medial frontal (MF), (5) Motor, (6) Salience (SN), (7) Subcortical (SBN), (8) Visual Association, (9) Visual I, (10) Visual II (Fig.2.A.). Lastly, the Subcortical network was divided into the Caudate (R/L), Thalamus (R/L), Hippocampus (R/L), Brainstem (R/L), Parahippocampus (R/L), and Putamen (R/L) for further investigation, as per ^46^. Due to the small size of the nodes in the last three regions, their R/L hemispheres were combined to form one univariate structure.

### Statistical analysis

The statistical analysis and data visualization were performed in Matlab and R. To calculate the FC matrices, the mean time-courses of every node-pair were Pearson correlated and their coefficients - Z-Fisher transformed ^46, 47^. This resulted in 35,778 unique edges for the whole brain of each subject, or 35,245 edges, if taking into account the removal of two “signal-loss” nodes. Functional connectivity for each network of the Shen268 atlas ^48^ was defined by averaging the connectivity between all pairs of nodes belonging to a specific network ^12^. The healthy controls (n = 113) were scanned on a comparable, but separate 3T Siemens machine and thus were used for visualization purposes only.

Next, Spearman’s correlation and R² analyses investigated any association between TBI patients’ Z-scores for each resting-state network and patients’ clinical measures (i.e. GCS or mRS) and the fit of the model, respectively.

The Mann-Whitney U test was used to establish any association between the connectivity Z-scores of the resting-state networks and patients’ GCS severity for the total GCS score and the subscores of the test (i.e. Eyes, Motor and Verbal). In this study, the GCS subscores were each binarized as follows: (1) Eyes “1-2” vs “3-4”, (2) Motor “1-5” vs “6” and (3) Verbal “1-4” vs “5”. Finally, GCS sub-group differences were evaluated within the Subcortical network using the same analytic approach. Associated p-values were Bonferroni corrected, except as indicated.

In this analysis, the small sample size did not allow for ordinal logistic regression or a higher classification model for the prediction of outcome based on acute/sub-acute data. Thus, for mRS at 3-months, we performed again the Mann-Whitney U test after binarizing the mRS values (i.e. mRS scores from “0-1” vs “2-6”/alternatively, mRS from “0-2” vs “3-6”)^49^. Associated p-values were Bonferroni corrected, except as indicated. Spearman correlation analysis was performed between the DMN FC scores and the mRS at 3-months.

Further, because of the non-normal distribution of the data, Kruskal-Wallis and Dunn’s analyses were conducted to explore the Subcortical FC across time and frequency (i.e. acute/longitudinal scan and/or first/repeat TBI). All associated p-values were Bonferroni corrected.

Lastly, ANCOVA was performed to test for the effect of motion noise and age on the resting-state networks. To quantify the magnitude of the reported results, partial eta squared was provided for the F-tests.

Considering the ongoing controversy regarding the use of the global signal regression (GSR) in resting-state analysis ^50^, the statistical analysis for the Shen268 atlas ^46^ was repeated using matrices that were pre-processed without the use of GSR. The data showed similar results irrespective of the use of GSR.

## Results

We screened 1,053 patients from the Yale New Haven Hospital database and identified 58 patients (mean ± SD, 41.28 ± 18.63years, 33% F) who met the inclusion criteria for our main analyses (Fig.1). An additional 50 patients (either scanned acutely/subacutely or longitudinally and with either a first or repeat TBI) were identified and combined with the main cohort of 58 patients for a post-hoc analysis, yielding altogether 108 patients (Fig.1.B). Then, we selected 113 healthy subjects for non-statistical (i.e. figure reference) comparison only (mean ± SD, 32.5 ± 12.6years, 52% F).

### Comparison of resting-state FC based on GCS (n = 58)

All main cohort patients combined (n = 58, Fig.1.A.) showed a significant Spearman’s correlation between Total GCS score and FC Z-scores of the (1) Subcortical (rho = 0.47, *P* < 0.001), (2) Motor (rho = 0.36, *P* = 0.005), (3) Salience (rho = 0.3, *P* = 0.021) and (4) DMN (rho = 0.33, *P* = 0.011). A subsequent coefficient of determination analysis showed R² = 0.18 (Subcortical), R² = 0.12 (Motor), R² = 0.15 (Salience) and R² = 0.09 (DMN), indicating that despite the significant correlation, GCS could not completely account for the variability in the FC of these resting-state networks and suggesting other factors might still contribute to this variability. Due to the clinical interest in the subscores of GCS, we also explored resting-state network relations with the Eye, Motor and Verbal subscores. With a single exception, these significant Spearman’s correlations for the Total GCS score were also confirmed across the GCS subscores (Supplementary Table 1).

Next, we dichotomized patients based on GCS severity (moderate-severe vs mild). The Mann-Whitney U test was performed to compare the difference in Z-scores FC between TBI groups, which were defined based on GCS Total score (moderate-severe vs mild) or the Eyes/Motor/Verbal GCS subscores. For the Total GCS score, we found statistically significant FC in the Subcortical network (U = 694, *P* < 0.001), Motor (U = 571, *P* = 0.018) and Salience networks (U = 575, *P* = 0.015), but not the DMN (U = 514, *P* = 0.147) (Fig.2.B). However, after multiple comparisons, only the Subcortical network remained statistically significant, with mild TBI patients showing higher median Z-scores than moderate-severe patients (Hodges-Lehmann (HL) estimate = 0.05, 95% CI [0.028, 0.072]).

For the individual GCS subscore analysis (i.e. Eyes/Motor/Verbal) (Fig.2.C), the Subcortical, Salience, Motor and DMN networks were found to statistically discriminate between subscore groups. However, after Bonferroni corrections, again only the Subcortical network remained statistically significant for all subscores (with *P* < 0.001) for Eyes (U = 209; HL = −0.037, 95% CI for HL [-0.062, −0.015]), Motor (U = 185; HL = −0.043, 95% CI [-0.064, −0.02]) and Verbal (U = 180; HL = −0.045, 95% CI [-0.065, −0.021]), with some exceptions for the Verbal subscore (Supplementary Table 2).

### Sub-Regions of the Subcortical resting-state network (n = 58)

Given the prominence of the Subcortical network in the GCS-based analysis, we conducted a subregional analysis by subdividing this network into its components based on the Shen268 atlas (Fig.3).

**Figure 3.**
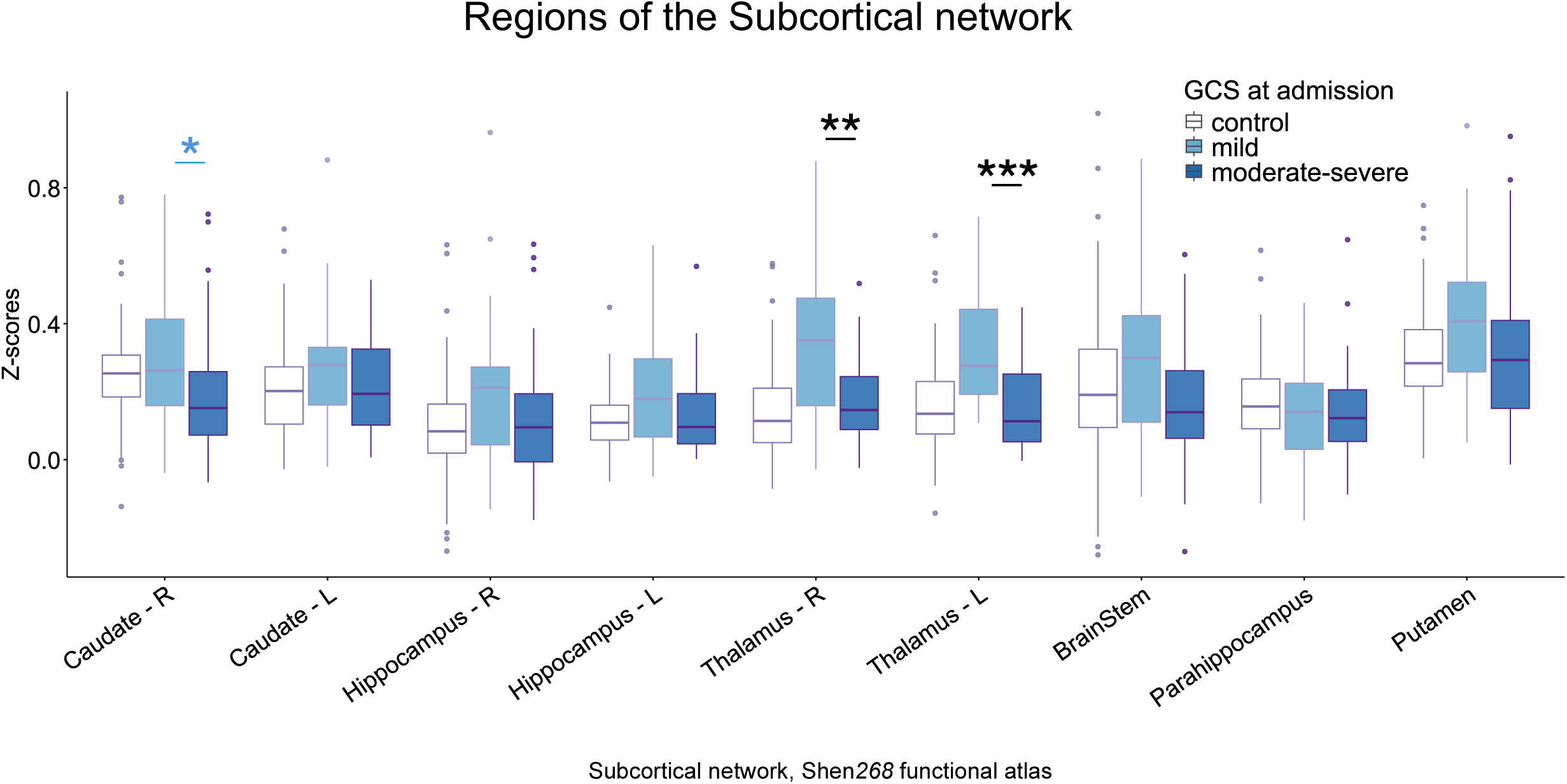
Glasgow Coma Scale (GCS) and functional connectivity alterations within the Subcortical network. The Mann-Whitney U test (total sample size n = 58, divided into n(mild) = 28 and n(moderate-severe) = 30) was performed to establish any association between the functional connectivity Z-scores within the Subcortical network sub-regions and patients’ GCS severity (i.e. moderate-severe vs mild). On the x-axis, we display the sub-regions of the Subcortical network, based on the Shen*268* functional atlas: caudate, hippocampus, thalamus, brainstem, parahippocampus and putamen. Due to the small size of the nodes in the last three regions (i.e. brainstem, parahippocampus and putamen), their R/L hemispheres were combined to form one univariate structure. On the y-axis, we display the functional connectivity Z-scores. Healthy controls (white, n = 113) are displayed for visual reference only. Each single point on the figure represents the Z-score of an individual participant’s functional connectivity map (i.e. patient or control) for the components of the Subcortical resting state network. **<u>Key:</u>** L – left, R – right, GCS – Glasgow Coma Scale; blue * denotes significant results that did not survive multiple comparisons; black * are results that remain statistically significant after Bonferroni corrections, * *p* < 0.05, ** *p* < 0.01, *** *p* < 0.001.

The Mann-Whitney U test indicated the Thalamus Right (U = 615, *P* = 0.002) and Thalamus Left (U = 661, *P* < 0.001) alongside the Caudate Right (U = 575, *P* = 0.015) as regions statistically discriminating between GCS severity groups. However, after multiple comparisons correction, only the Thalamus (R/L) remained statistically significant (Right - (HL = 0.178, 95% CI [0.063, 0.289]), Left - (HL = 0.16, 95% CI [0.083, 0.237])).

### mRS at 3months and its prediction power based on resting-state FC (n = 58)

Spearman correlation analysis between mRS at 3-months and the FC Z-scores of the 10 resting-state networks showed significant associations to the (1) DMN (rho = −0.382, *P* = 0.003) and (2) Motor (rho = - 0.365, *P* = 0.005) networks. A coefficient of determination analysis showed moderate R² = 0.18 (DMN) (Supplementary Fig.2) and R² = 0.12 (Motor), indicating that despite the strong statistical correlation to these two resting-state networks, mRS at 3-months could not explain a large portion of the variability in the neural correlates represented by the DMN/Motor resting-state FC.

Given inconsistencies in the literature defining optimal cutoff points for dichotomizing functional outcome scales, we evaluated resting-state network differences using two mRS groupings for “minor symptoms” vs “symptomatic (0-1 vs 2-6 and 0-2 vs 3-6). Using these two binarization methods, we performed the Mann – Whitney U test across the 10 resting-state networks (Fig.4.A). These comparisons showed that the 0-2 vs 3-6 division better aligned with the correlation analyses compared to the 0-1 vs 2-6 division, particularly for the DMN. While the 0-1 vs 2-6 mRS division only showed a trend towards significance in the DMN (U = 512, *P* = 0.083), the mRS 0-2 vs mRS 3-6 division showed a significant difference in the DMN (U = 527, *P* = 0.016, HL = 0.051, 95% CI [0.008, 0.091]) (Fig.4.A). The latter analysis suggested the difference might be driven by patients with an mRS2 (n = 15). However, these patients had an evenly distributed GCS score (seven: - GCS 6-8; and eight: GCS 14-15). Additionally, even after excluding patients with mRS2, the difference in DMN FC remained statistically significant.

**Figure 4.**
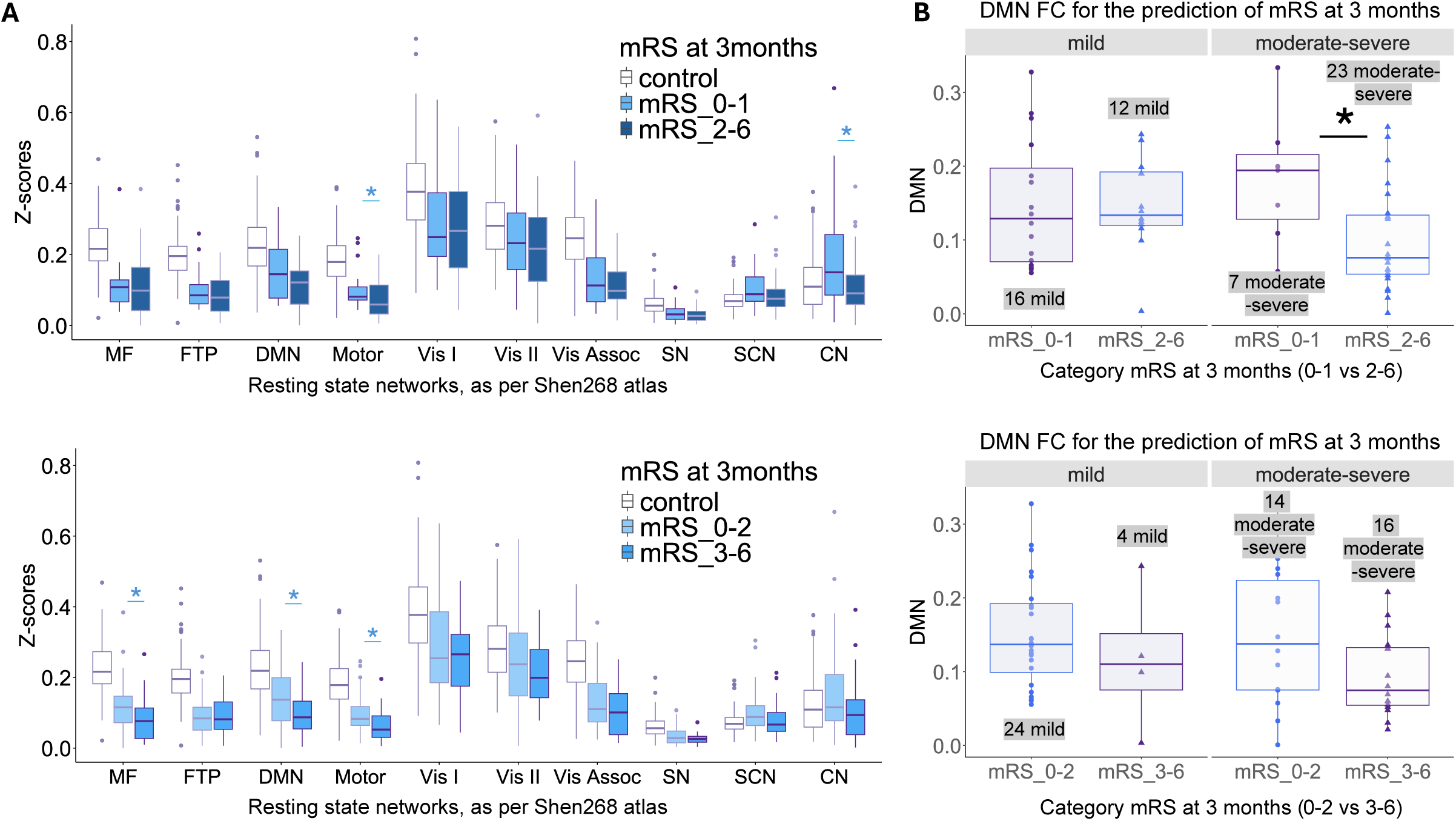
Associations between functional connectivity (FC) patterns and modified Rankin Scale (mRS) at 3months. **(A).** Network analysis based on mRS at 3 months. Outcomes were dichotomized based on two divisions: (1) minimal (mRS 0-1) vs persistent/disabling symptoms (mRS 2-6); (2) independent (mRS 0-2) vs moderate/severe symptoms (mRS 3-6). The Mann-Whitney U test (total sample size n = 58, divided into either n[mRS (0-1)] = 23 and n[mRS (2-6)] = 35 (panel above) or alternatively into n[mRS (0-2)] = 38 and n[mRS (3-6)] = 20 (panel below)) was used to establish any association between the functional connectivity Z-scores of the resting-state networks and patients’ mRS scores at 3months. The x-axis shows 10 resting-state networks based on the Shen*268* functional atlas, and the y-axis displays the Z-scores of the functional connectivity. Healthy controls (white, n = 113) are displayed for visual reference only. Each single point on the figure represents the Z-score of an individual participant’s functional connectivity map (i.e. patient or control) for the corresponding resting state network. **<u>(B).</u>** DMN functional connectivity for the prediction of mRS at 3 months in TBI patients based on GCS at admission. Patients are first split into two plots based on mRS dichotomization along the x-axis (either 0-1 vs 2-6 (left plot) or 0-2 vs 3-6 (right plot)). The y-axis shows the Z-score functional connectivity of the DMN. Each plot is then further divided based on TBI severity, classified as “mild” or “moderate-severe” using total GCS on admission. The Mann-Whitney U test was used to compare between mRS subgroups, e.g., between 0-1 vs 2-6 within the moderate-severe GCS group. No multiple comparison test was required for this part of the analysis. Each single point on the figure represents the Z-score of an individual patient’s functional connectivity map for the DMN. **Sample sizes** - <u>left plot, left panel</u> – mild TBI GCS patients divided into their mRS scores – 0-1 (n = 16) and 2-6 (n = 12), <u>left</u> <u>plot, right panel</u> – moderate-severe TBI GCS patients divided into their mRS scores – 0-1 (n = 7) and 2-6 (n = 23), <u>right plot, left panel</u> – mild TBI GCS patients divided into their mRS scores – 0-2 (n = 24) and 3-6 (n = 4), <u>right plot, right panel</u> – moderate-severe TBI GCS patients divied into their mRS scores – 0-2 (n = 14) and 3-6 (n = 16). **<u>Key:</u>** blue * denote significant results that did not survive multiple comparisons; black * are results that remain statistically significant after Bonferroni corrections, * *p* < 0.05, ** *p* < 0.01, *** *p* < 0.001; Medial-Frontal network = MF, Fronto-Parietal network = FTP; Default Mode Network = DMN, Visual I network = Vis I, Visual II network = Vis II, Visual Association = Vis Assoc, Salience = SN, Subcortical network = SCN, Cerebellar network = CN.

Beyond the DMN, the 0-1 vs 2-6 dichotomization also revealed statistical discrimination between patients’ scores at 3-months in the (1) Motor (U = 536, *P* = 0.034, HL = 0.028, 95% CI [0.004, 0.048]) and (2) Cerebellum networks (U = 530, *P* = 0.043, HL = 0.055, 95% CI [0.0005, 0.12]). On the other hand, the alternative 0-2 vs 3-6 division indicated the importance of the (1) Motor network (U = 519, *P* = 0.023, HL = 0.029, 95% CI [0.004, 0.052]) and (2) the Medial - Frontal network (U = 518, *P* = 0.024, HL = 0.037, 95% CI [0.007, 0.077]), alongside the DMN results.

None of the mRS analysis results, irrespective of the mRS dichotomization used, survived multiple comparison corrections (Fig.4.A).

### Prediction of GCS at admission to mRS at 3-months (n = 58)

Next, we explored whether admission GCS could predict 3-month functional outcome based on DMN connectivity during the acute/subacute timeframe (Fig.4.B).

Regardless of how the mRS was dichotomized, DMN FC did not predict functional outcome based on mRS at 3-months in mild TBI patients (Fig.4.B - right plot, left panel; Fig.4.B - left plot, left panel). However, when evaluating moderate-severe TBI patients, DMN Z-scores managed to predict the functional outcome based on 3-month mRS when comparing 0-1 vs 2-6 as assessed by the Mann-Whitney U test (Fig.4.B - left plot, right panel) (U = 123, *P* = 0.037, HL = 0.078, 95% CI [0.01, 0.152]). A similar trend was observed when comparing mRS 0-2 vs mRS 3-6 with higher DMN Z-scores in the better outcome group, although this did not reach statistical significance (Fig.4.B - right plot, right panel) (U = 150, *P* = 0.12). This lack of significance may be partially explained by the inclusion of several patients (n=7) in the mRS 2 group, all of whom had severe GCS scores.

Of note, only mild and moderate patients achieved “complete” recovery at 3-month post-TBI, based on their mRS = 0 at 3-months (i.e. 9% if only “mRS 0”, or 39% if “mRS 0 + mRS 1”).

### Subcortical FC from acute to longitudinal and from first to repeat TBI (n = 108)

A post-hoc analysis was performed to further explore Subcortical FC over time, frequency and severity of TBI (Fig.5). Thus, we extended our cohort (Fig.1.B) with patients who were scanned outside of the acute window (i.e. had a “longitudinal scan”) or had a history of a prior TBI (i.e. had a “repeat TBI”). Our extended cohort of 108 patients was divided into four groups based on the time between the traumatic event and the scan, alongside frequency of TBI (i.e. first/repeat). Thus, we had (1) acute/sub-acute first TBI (n = 58), (2) acute/sub-acute repeat TBI (n = 12), (3) longitudinal first TBI (n = 28) and (4) longitudinal repeat TBI (n = 10). The acute/sub-acute first TBI group continued to be separated into mild and moderate-severe patients (28 mild + 30 moderate-severe, median group GCS = 15 and GCS = 7, respectively). All other groups were kept as a single group due to their smaller sample size and predominant GCS scores. In particular, (A) acute/sub-acute repeat TBI (4 mild + 8 moderate-severe, median group GCS = 9), (B) longitudinal first TBI (25 mild + 3 moderate – severe, median group GCS = 15) and (C) longitudinal repeat TBI (8 mild + 2 moderate-severe, median group GCS = 15).

**Figure 5.**
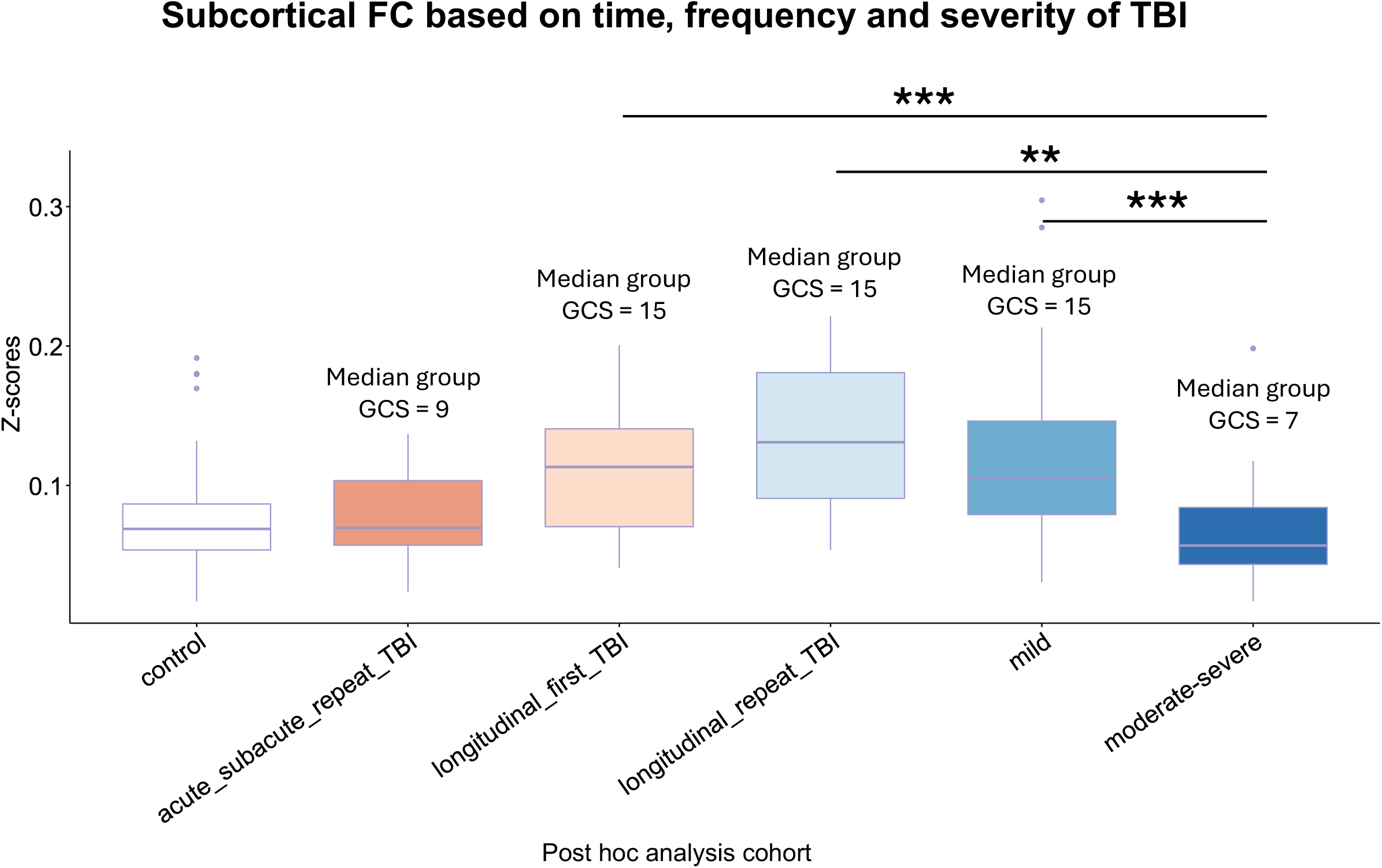
Subcortical functional connectivity (FC) based on time, frequency and severity of TBI. Differences in functional connectivity of the Subcortical network based on TBI severity (mild vs moderate-severe), TBI acuity of scan acquisition (acute or longitudinal) and exposure to TBI (acute or repeat injury) are displayed. The x-axis shows five distinct patient groups that constitute the post hoc analysis: **(1)** acute/sub-acute mild first TBI (n=28), **(2)** acute/sub-acute moderate-severe first TBI (n=30), **(3)** acute/sub-acute repeat TBI (n=12), **(4)** longitudinal first TBI (n=28), **(5)** longitudinal repeat TBI (n=10). The y-axis displays the Z-scores of the Subcortical functional connectivity. Above each boxplot, we report the median group GCS representative of the specific patient group. Healthy controls (n = 113) are displayed for visual reference only. The Kruskal- Wallis test assessed Subcortical functional connectivity differences across severity, time and frequency. Dunn’s test with Bonferroni correction identified pairwise comparisons remaining significant after multiple comparisons. Each single point on the figure represents the Z-score of an individual participant’s functional connectivity map (i.e. patient or control) for the Subcortical resting state network. **<u>Key:</u>** acute/subacute scan (i.e. <= 31 days); longitudinal scan (>31 days post injury up to 2 years); repeat (>1 TBI event); * *p* < 0.05, ** *p* < 0.01, *** *p* < 0.001.

The Kruskal- Wallis for Subcortical FC across time and frequency range was significant (χ2(4) = 28.31, *P* < 0.001, η_p_^2^ = 0.24). Subsequent Dunn’s *post hoc* confirmed a notable effect of sub-group for (A) mild and moderate-severe TBI (acute/sub-acute first TBI) (*P_bonf_* < 0.001), (B) moderate-severe TBI (acute/sub-acute first TBI) and longitudinal first TBI (*P_bonf_* = 0.001) and (C) moderate-severe TBI (acute/sub-acute first TBI) and longitudinal repeat TBI (*P_bonf_* = 0.002).

We also explored the link between the Thalamus and the rest of the resting-state networks not only during the acute/sub-acute stage, but also over time and frequency of TBI (Supplementary Fig.1).

### Motion noise and Age as covariates

Motion noise is an essential concern in MRI scans, even more so in clinical scans. Mean motion noise for each patient was calculated as the mean of the frame-to-frame displacement of the functional image < 1.2mm. After a robust motion noise cleaning procedure, motion noise from the clinical scans still correlated to Total GCS (*P* < 0.001) and the Subcortical network (*P* < 0.001), but not to the DMN (*P* = 0.83) or mRS (*P* = 0.25). After regressing out the motion noise [F(1,54) = 18.21, *P* < 0.001, η_p_ ^2^ = 0.25], ANCOVA showed that GCS still retained its meaningful impact on the discrimination of Subcortical FC Z-scores among patients [F(1,54) = 24.13, *P* < 0.001, η_p_ ^2^ = 0.31].

Similarly, we accounted for the effect of age – age was not notably correlated to any resting-state network or mRS, but to GCS (*P* < 0.001). However, ANCOVA confirmed that age did not affect [F(1,54) = 0.035, *P* = 0.85, η_p_ ^2^ < 0.001] the discrimination of mild/moderate-severe patients within the Subcortical network.

Importantly, in this study, we did not use GCS Total as a continuous variable (which initially showed the significant correlation), but a dichotomized variable based on the continuous one. Thus, the above-mentioned considerations might be unnecessary.

## Discussion

In this study, we investigated the relationship between resting-state FC and two common clinical measures, based on TBI severity and functional outcome. We found that acquiring acute/sub-acute FC scans may provide complementary functional brain-based information about TBI severity to GCS by highlighting the utility of resting-state substrates, particularly of the Subcortical network. In addition, our findings suggest the DMN may correlate with functional outcome at 3-months post-TBI in moderate-severe TBI patients.

### GCS at admission was reflected in the Subcortical network

Ideally, a clinical score should be a relatively good representation of the canonical brain networks to ensure that disruptions caused by injuries like TBI are accurately captured in routinely collected clinical assessments. However, based on our findings and previous research ^2, 3, 4, 5^, the GCS appears to lack the sensitivity to broadly reflect these network perturbations. While the total GCS was significantly linked to the Subcortical, DMN, Salience and Motor networks based on our correlation analysis, the low R^2^ values suggested that GCS failed to explain a lot of the variability in these networks. Many TBI studies focus on one specific severity sub-group in their analyses ^20, 22, 16^, thereby limiting our understanding of TBI brain-behavior correlates when trying to compare across the full spectrum of TBI severity. While there is certain natural variability that is not related to brain injuries, clinical tools most commonly used to assess TBI severity, like GCS, remain cursory.

A key finding of this study was the limited capture of GCS when constructing a comprehensive brain-behavioral representation of TBI severity. Nevertheless, after examination of the 10 resting-state networks ^46, 47^ the Subcortical network discriminated significantly between mild/moderate-severe TBI patients. Since GCS primarily reflects patient consciousness levels, our finding that its discriminative power was largely centered in the Subcortical network during acute/sub-acute TBI is well supported ^51, 52, 53^. This observation was independent of whether the investigation was done using Total GCS or GCS Subscores (i.e. Eyes, Motor, Verbal). Previous research stressed the importance of this network in showing its abnormal FC in disorder of consciousness patients ^51, 52, 53^. Together with the DMN and other cortical networks, the Subcortical network indicated that its degree of alteration might be linked to the level of consciousness impairment and that subsequent recovery was a consequence of the normalization of its connectivity ^52^.

Despite its central role in the establishment of consciousness ^54, 31^, in our study, DMN connectivity did not survive correction for multiple comparisons when discriminating between mild/moderate-severe GCS during the acute/sub-acute phase. This suggests that GCS is either too superficial to distinguish between TBI categories in other canonical networks, e.g. the DMN, or that these networks can only discern a pattern in a patient-control, but not in the subtler patient-patient interaction.

### Mild TBI hyperconnectivity vs moderate-severe TBI hypoconnectivity

Further analysis among the regions of the Subcortical network pointed to the right and left thalami, substantiating not only the potential importance of the brain’s relay station for mild TBI and patients’ outcome ^55, 56, 57^ but also for the initial response to brain trauma ^58,59^ and consciousness ^51, 60^. Interestingly, the thalamus not only showed the only statistically different FC levels among GCS severity groups, but its strongest FC was within the Subcortical network, rather than outside of this network (Supplementary Fig.1).

Additionally, we found that at both Subcortical and thalamic levels, mild TBI patients exhibit expressed hyperconnectivity relative to their moderate-severe TBI counterparts. This suggests the two severity groups may not rely on the same compensatory mechanisms post-injury. Hyperconnectivity is suggested to be a basic response to neurological disease, including TBI ^61, 62, 63^. It has mainly been proposed as a response to enhance the neural network performance and restore communication, recruiting increased resources to detour otherwise damaged pathways. Both centralized and regional hyperconnectivity have previously been described. Regional hyperconnectivity depends on the use of regional resources, whereas centralized hyperconnectivity uses well-established critical network hubs (one of which is the thalamus) and does not depend on the etiology or location of the TBI ^62^. When compared, centralized hyperconnectivity, which reflects a group of highly connected and conveniently positioned nodes ^62^, is more metabolically efficient than regional increases spread across the brain. Our findings of thalamic hyperconnectivity in mild TBI are corroborated by previous research ^19^, and support a potential theory of bypassing damaged pathways by recruiting centralized thalamic networks in search of compensatory mechanisms.

Interestingly, functional hyperconnectivity in TBI is a more often cited response to TBI than hypoconnectivity, the latter considered to be linked to extensive resource loss in the affected regions ^61^, as seen in severe TBI cases like our moderate-severe cohort. By being able to make a direct comparison across TBI severities, the network pathology among TBI patients appeared exaggerated within the specified region examined. We postulate two reasons for the observed Subcortical hypoconnectivity: (1) that overall damage was too critical and thus centralized attempts to compensate were not possible, resulting in persistently low FC levels, (2) moderate-severe TBI brain mechanisms attempted to offload to the thalamus, but the overstimulation was too energy-consuming, leading to metabolic exhaustion and relative hypointensity.

Lastly, the MRI radiological reports of the acute/sub-acute patients did not mention structural damage to the thalamus. This was detected by the increased sensitivity of the resting-state functional maps.

### At 3-months, mRS showed a correlation to the DMN resting-state FC

To evaluate patients’ outcome at 3-months, we examined FC Z-scores in the 10 canonical resting-state networks explored in this study ^46,47^. The mRS provides a global measure of the cumulative impact of large-scale network disruptions, reflecting the integrated effects of impaired mobility, slowed processing, and increased dependency. Thus, mRS serves as a clinically meaningful anchor for interpreting rsfMRI findings, linking network-level physiology to real-world independence and disability.

When comparing patients with “no/minor symptoms” (mRS 0-1) versus “moderate/severe symptoms” (mRS 2-6), we found that the differences mainly emphasized networks related to motor functions (i.e. the Motor network) and balance (i.e. the Cerebellum network). On the other hand, dichotomizing patients based on the likelihood of functional and cognitive dependence (mRS 0-2 vs mRS 3-6) highlighted differences in the Medial-Frontal network and DMN. Still, we remain cautious about the overinterpretation of these results, as neither of them remained statistically important after Bonferroni corrections. Whereas there are more task-based fMRI studies which support fMRI prediction of TBI outcome ^52, 64^ fewer resting-state studies were conducted in the acute state post-TBI. However, those that exist appear to support a correlation between reduced DMN FC and functional outcome ^65^, though predictive performance remains ambiguous ^30^. Interestingly, the Subcortical network did not statistically discriminate between patient outcomes at 3-months. This may be related to the limited resolution of mRS and/or to the broader symptoms and functions assessed for when calculating mRS compared to GCS, thus no longer highlighting the contrast in TBI GCS severity noted previously in the Subcortical network.

### Subcortical FC appeared to be reflective of the severity of the injury

In our final post-hoc analysis, we aimed to understand whether chronicity, frequency of injury or injury severity has the greatest impact on hyperconnectivity of mild TBI patients and/or the hypoconnectivity of moderate-severe TBI observed in the Subcortical network. Despite the complexity of interpreting repeat TBI and the lack of understanding of its molecular mechanisms ^66^, these patients constitute a commonly presenting population of TBI patients to the ER. Therefore, considering that they are more susceptible to neurodegeneration ^67, 66, 68^, more research is warranted to elucidate the neuronal sequelae of their TBI. Our results suggested that in our cohort, injury severity, as measured by the GCS, had a more pronounced effect on FC than chronicity or frequency of injury. However, combining mild injury severity with repeated injury appeared to produce the most pronounced hyperconnectivity in mild TBI patients compared to all other groups. Previous research comparing first vs repeat mild TBI (scanned on average 12.5days (SD ± 9.9) post TBI) ^68^ showed that the repeat TBI patients had an even further exacerbated hyperconnectivity compared to the acute first TBI, postulating the neurological effects of repeat TBI might be cumulative ^68^. While differentiation among mild TBI patients might highlight injury frequency as a significant factor, the inclusion of a moderate-severe TBI cohort in our study revealed that TBI GCS severity appears to be the primary determinant of FC differences among groups. Still, we acknowledge that GCS has important limitations as a sole TBI severity index. Thus, we included it mainly as it is a common clinical benchmark that enables contextualizing rsfMRI findings and comparison to prior literature. Further, as an advanced imaging technique, rsfMRI may not yet be well adapted into the clinical workflow, but it represents an alternative for severe TBI patients unable to undergo a task-fMRI. Thus, this clinical protocol attempted to make fMRI more broadly available to all TBI patients.

While our and previous research ^19, 68^ suggests that Subcortical hyperconnectivity persists over time, it is unclear how advantageous such a response might be long-term. While undoubtedly beneficial in the short-term, long-term thalamic hyperconnectivity may lead to the network hubs becoming more susceptible to secondary pathological processes due to “chronically elevated metabolic stress” ^62^. Research suggests that long-term hub “overload” might be a sign of pathology ^69^, and these sites are vulnerable to increased amyloid beta deposition, a marker of Alzheimer’s disease ^62^. As there may be an ultimate “cost-efficiency negotiation” during the follow-up period as a response to the brain optimization and adaptation to the energy-consuming demands of the injury ^63, 62^, all this might hold numerous implications for long-term outcome after TBI ^63^.

### Limitations of this study

This single-center study provided a rare opportunity to examine ER patients using rsfMRI. Still, despite its strengths, this study has several limitations. First, collecting acute/sub-acute MRI scans remains challenging resulting in small sample sizes. Second, healthy participants could not be scanned on ER scanners, which made us introduce a group of institutional proxy controls, representative of the average healthy performance. Still, this was done for visualization purposes only to avoid over-inflation of the data. Importantly, this kept the focus on the difference between patient groups, that despite being more subtle, is more clinically relevant to our question. Third, the thalamus was not segmented into nuclei as there is currently no widely accepted thalamic functional atlas, despite several valuable examples ^70^. Fourth, our functional outcome scores were limited to the 3-month follow-up, suggesting that longer timeframes are needed to reassess the correlations and prediction analyses. Lastly, this study is reflective of real-world ER data, where patients’ notes and time for examination are limited. Although more detailed assessments could be used in a structured research setting, such an approach would not capture the practical constraints of the metrics guiding initial ICU diagnosis.

## Conclusion

Our goal in this study was to determine whether two widely used clinical tools, GCS and mRS, could be used to link to the brain’s performance and diagnostic prognostication after TBI. We found that GCS at admission is reflected by increased resting-state FC in the Subcortical network, particularly in the thalamus. This increased FC showed clear discrimination between GCS severity groups, with hyperconnectivity in mild TBI patients. Our study also showed that the DMN FC in the acute/sub-acute stage showed a potential link to 3-month mRS performance, which could provide valuable prognostication if confirmed in a larger cohort.

Despite some meaningful insights, GCS and mRS showed limited sensitivity to the TBI brain-behavior representation, suggesting that alternative clinical assessments may better reflect early pathological connectivity. Consistent with recent evidence, these results suggest that GCS alone can inadequately capture acute TBI severity, whereas FC offers complementary prognostic insight.

## Data availability

Anonymized raw data is available upon reasonable request.

## Acknowledgements

The authors would like to thank all patients whose records were used in this retrospective review study.

## Supplementary

**Supplementary Table 1.**
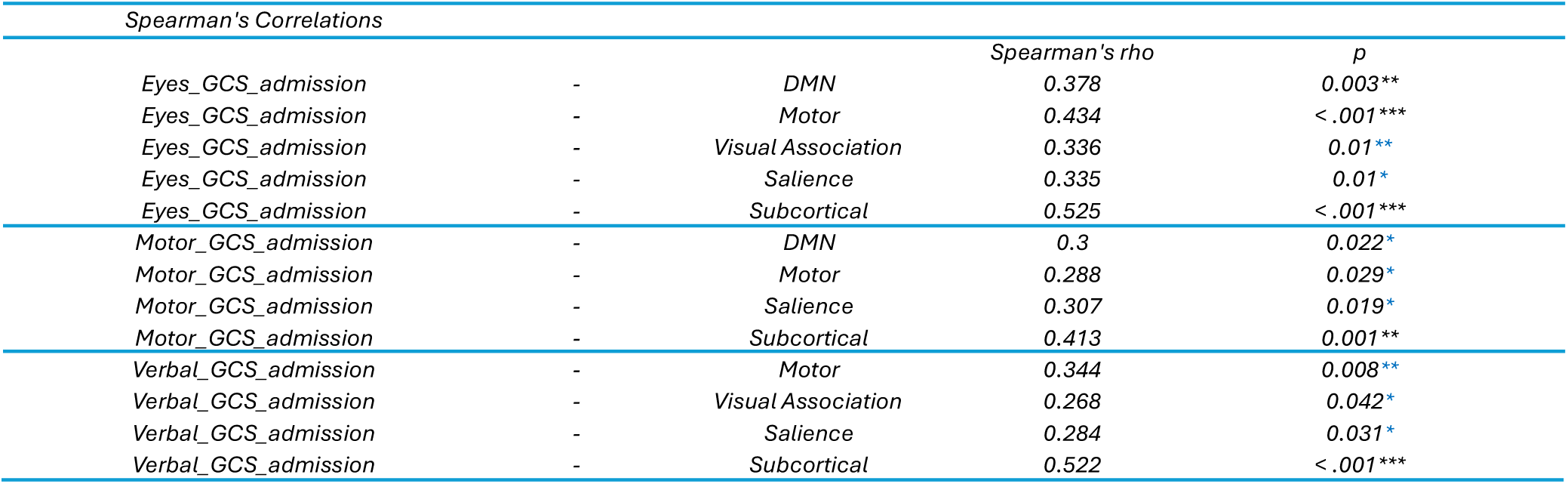
Spearman correlations between GCS subscores and resting-state networks of all TBI patients. Due to space constraints, only statistically significant correlations (before or after multiple comparisons) are displayed. **<u>Key</u>:** blue * denotes significant results that did not survive multiple comparisons; black * are results that remain statistically significant also after Bonferroni corrections, * *p* < 0.05, ** *p* < 0.01, *** *p* < 0.001.

| Spearman's Correlations |  |  | Spearman's rho | p |
| --- | --- | --- | --- | --- |
| Eyes_GCS_admission | - | DMN | 0.378 | 0.003** |
| Eyes_GCS_admission | - | Motor | 0.434 | < .001*** |
| Eyes_GCS_admission | - | Visual Association | 0.336 | 0.01** |
| Eyes_GCS_admission | - | Salience | 0.335 | 0.01* |
| Eyes_GCS_admission | - | Subcortical | 0.525 | < .001*** |
| Motor_GCS_admission | - | DMN | 0.3 | 0.022* |
| Motor_GCS_admission | - | Motor | 0.288 | 0.029* |
| Motor_GCS_admission | - | Salience | 0.307 | 0.019* |
| Motor_GCS_admission | - | Subcortical | 0.413 | 0.001** |
| Verbal_GCS_admission | - | Motor | 0.344 | 0.008** |
| Verbal_GCS_admission | - | Visual Association | 0.268 | 0.042* |
| Verbal_GCS_admission | - | Salience | 0.284 | 0.031* |
| Verbal_GCS_admission | - | Subcortical | 0.522 | < .001*** |

**Supplementary Table 2.** Discrimination between GCS subscores groups based on their functional connectivity. **<u>Key:</u>** blue * denotes significant results that did not survive multiple comparisons; black * are results that remain statistically significant after Bonferroni corrections, * *p* < 0.05, ** *p* < 0.01, *** *p* < 0.001.

| Eyes-GCS at admission |  |  | Motor-GCS at admission |  |  | Verbal-GCS at admission |  |  |
| --- | --- | --- | --- | --- | --- | --- | --- | --- |
|  | U | P |  | U | P |  | U | P |
| Medial Frontal | 355 | 0.347 | Medial Frontal | 324 | 0.168 | Medial Frontal | 252 | 0.016* |
| Fronto-parietal | 359 | 0.38 | Fronto-parietal | 323 | 0.163 | Fronto-parietal | 254 | 0.018* |
| DMN | 284 | 0.039* | DMN | 278 | 0.035* | DMN | 273 | 0.04* |
| Motor | 266 | 0.019* | Motor | 256 | 0.013* | Motor | 212 | 0.002** |
| Visual I | 326 | 0.163 | Visual I | 292 | 0.059 | Visual I | 298 | 0.099 |
| Visual II | 433 | 0.798 | Visual II | 415 | 0.975 | Visual II | 311 | 0.149 |
| Visual Association | 304 | 0.081 | Visual Association | 325 | 0.173 | Visual Association | 219 | 0.003** |
| Salience | 258 | 0.013* | Salience | 271 | 0.026* | Salience | 246 | 0.012* |
| Subcortical | 209 | < .001*** | Subcortical | 185 | < .001*** | Subcortical | 180 | < .001*** |
| Cerebellum | 347 | 0.287 | Cerebellum | 316 | 0.132 | Cerebellum | 304 | 0.12 |

**Supplementary Figure 1.**
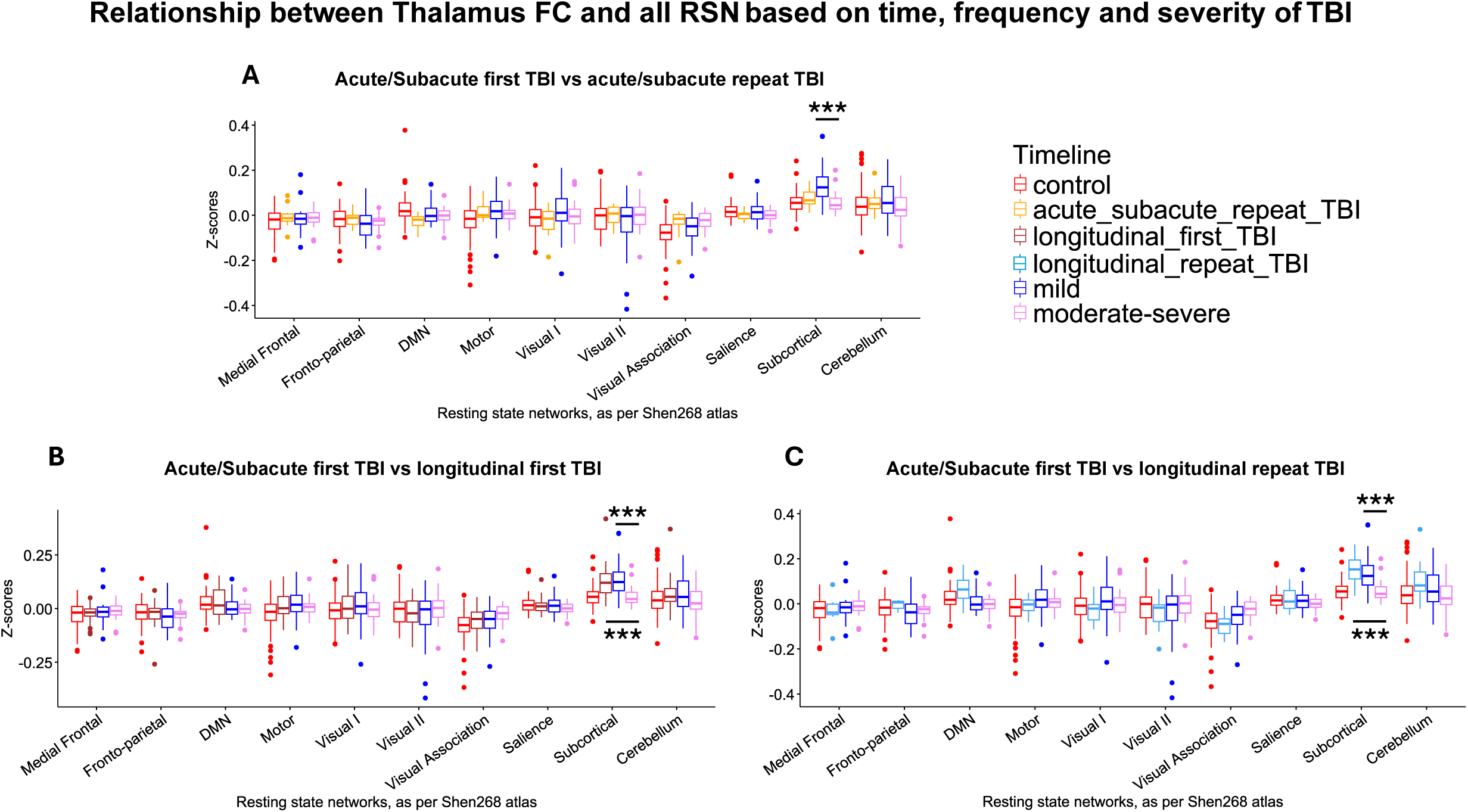
Relationship between Thalamic functional connectivity (FC) and all resting-state networks based on time, frequency and severity of TBI. A Kruskal- Wallis test was used for each individual subplot (A), (B) and (C) and assessed functional connectivity differences across severity, time and frequency. Dunn’s test with Bonferroni correction identified pairwise comparisons remaining significant after multiple comparisons. For instance, in (A), the Kruskal-Wallis test assessed the difference between (1) acute/subacute mild first patients, (2) acute/subacute moderate-severe first patients and (3) acute/subacute repeat TBI. As the Kruskal-Wallis test was statistically significant, we did a Dunn’s test identifying the statistical importance between acute/subacute first TBI patients (i.e. mild vs moderate-severe). The same logic applied to (B) and (C). Each single point represents the Z-score of an individual participant’s (i.e. patient or control) relationship between the thalamus and the corresponding resting state network, indicated on the x axis. **<u>(A). Acute/Subacute first TBI vs acute/subacute</u> <u>repeat TBI</u>** - Comparison between Thalamic acute/subacute first TBI and Thalamic acute/subacute repeat TBI functional connectivity. **<u>(B). Acute/Subacute first TBI vs longitudinal first TBI</u>** - Comparison between Thalamic acute/subacute first TBI and Thalamic longitudinal first TBI functional connectivity maps.**<u>(C). Acute/Subacute first TBI vs longitudinal repeat TBI</u>** - Comparison between Thalamic acute/subacute first TBI and Thalamic longitudinal repeat TBI patients. **<u>Key</u>:** blue * denote significant results that did not survive multiple comparisons; black * are results that remain statistically significant after Bonferroni corrections, * *p* < 0.05, ** *p* < 0.01, *** *p* < 0.001; on all plots – healthy controls (n = 113), acute/subacute mild first TBI (n = 28), acute/subacute moderate-severe first TBI (n = 30), acute/subacute repeat TBI (n = 12), longitudinal first TBI (n = 28) and longitudinal repeat TBI (n = 10).

**Supplementary Figure 2.**
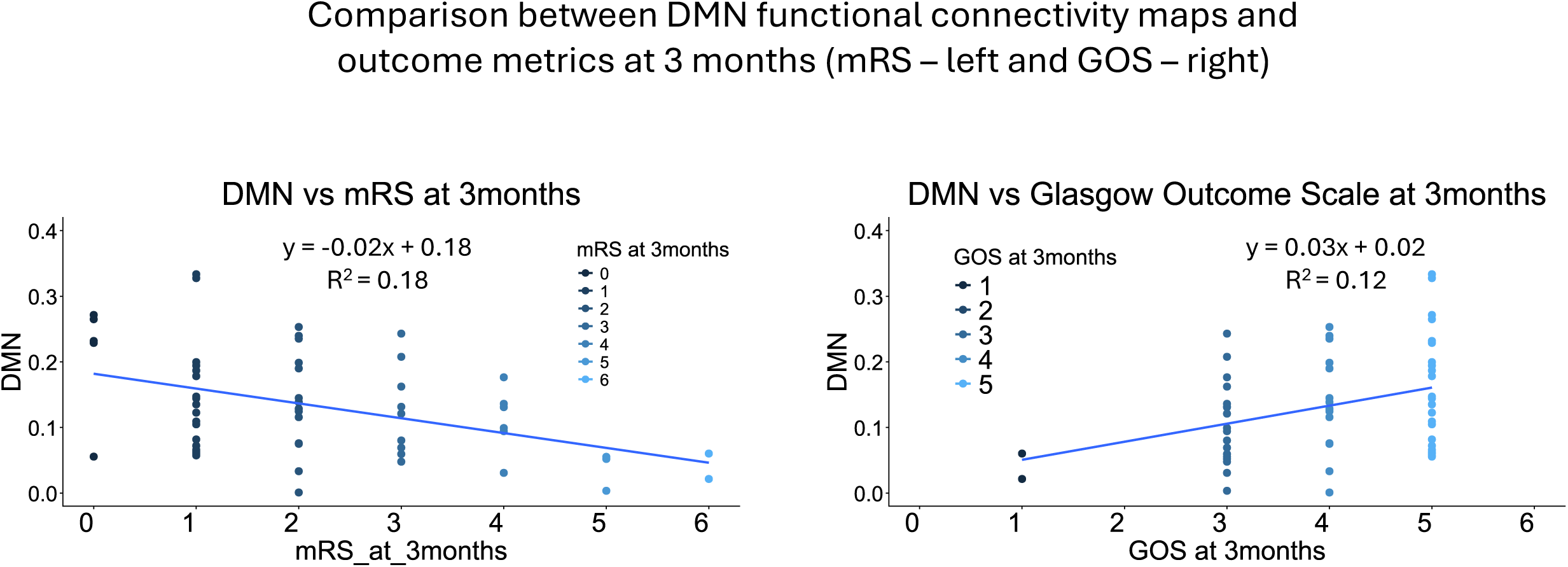
Comparison between Modified Rankin scale (mRS) and Glasgow Outcome scale (GOS). The left and right panel provide a comparison of the acute/subacute functional connecitivity of the DMN (n = 58, main cohort for our study) using either mRS (on the left, similar to our main analysis) or GOS (hypothetical suggestion) as the outcome measure. The x-axis displays the clinical metric (mRS on the left or GOS – on the right), whereas the y-axis displays the Z-score of the DMN functional connectivity. Each single point represents the Z-score of an individual patient’s functional connectivity map for the DMN. The R^2^ analysis and the slope show the fit of the regression model, indicating that there is a good correspondence between the 2 models as performance seems comparable. The transformation of the mRS score to GOS was done as per *Gaastra et al, 2022*. In particular, mRS 0 and mRS 1 = GOS 5, mRS 2 = GOS 4, mRS 3 and mRS 4 and mRS 5 = GOS 3 and finally mRS 6 = GOS 1. **Key**: DMN – default mode network, mRS – Modified Rankin scale, GOS – Glasgow Outcome scale.

